# The Effect of Obesity on Pain Severity and Pain Interference

**DOI:** 10.1101/2020.03.02.20027425

**Authors:** Jade Basem, Robert S. White, Stephanie A. Chen, Elizabeth Mauer, Michele L. Steinkamp, Charles E. Inturrisi, Lisa Witkin

## Abstract

**Background and objectives:** Obesity is one of the most prevalent comorbidities associated with chronic pain, the experience of which can severely interfere with activities of daily living and increase the utilization of clinical resources. Obesity is also a risk factor for increased pain severity (pain intensity) and pain interference (pain related disability). We hypothesize that a higher level of obesity, as measured by body mass index (BMI), would be associated with increased levels of pain severity and interference in a population of chronic pain clinic patients.

**Methods:** Participant data was pulled from a multi-site chronic pain outpatient database from 7/8/2011 to 10/17/2016. The Brief Pain Inventory (BPI), opioid prescriptions, and basic demographic information were queried and we categorized participants into three different ordinal categories based on recorded BMI levels (underweight, normal and overweight, obese). Bivariate analyses were performed to compare pain outcomes by BMI and by other demographic/clinical patient characteristics. Multivariable linear regressions were constructed to model each of four pain severity scores in addition to total pain interference score. All models examined BMI as the primary predictor, controlling for age, receipt of a pain procedure within 45 days prior to the pain clinic encounter, opioid prescription within 45 days prior to the encounter, and diagnosis. The total pain interference model additionally included pain severity (as measured by worst pain in the past 24 hours) as a covariate.

**Results:** 2509 patients were included in the study. The median BMI was 27 and the median age was 59 years. 77% of patients were diagnosed with musculoskeletal pain conditions. Bivariate tests revealed significant differences between BMI groups for all pain severity scores and for total pain interference score. On multivariable modelling controlling for age, pain procedure within 45 days prior to pain clinic encounter, opioid prescription with 45 days prior, and diagnosis, obese patients had significantly higher pain severity (as measured by worst, least, average, and current pain in the past 24 hours) as well as higher pain interference (as measured by the overall pain interference score) than normal weight and overweight patients.

**Conclusion:** In our study of pain clinic patients, obesity was found to be associated with increased pain severity and pain interference. We believe that this relationship is multifactorial and bidirectional. Pain phyisicans should consider the impact of obesity when addressing pain management for patients.

## INTRODUCTION

The Institute of Medicine estimates that 100 million United States adults are affected by chronic pain, which costs the nation $635 billion yearly in medical treatment and lost productivity ^1^. Pain is a common experience for the general population, as 11.2% of adults report daily pain symptoms ^2^. Pain is the most common reason patients seek medical attention with approximately one-fifth of ambulatory medical care associated with pain as a primary symptom or diagnosis ^3; 4^. The experience of pain is complex, as chronic pain is a multidimensional hierarchical construct that consists of both the sensation of pain severity and interference in engagement with activities^5^. In addition, a threshold effect occurs where low levels of pain intensity are not associated with functional interference; however, higher levels of pain intensity increase the probability of interference in a nonlinear fashion ^5-7^.

One of the prevalent comorbidities that is commonly associated with chronic pain is obesity ^8^. Data from the National Health and Nutrition Examination Survey from 2015-2016 demonstrates that 39.8% of United States adults were obese ^9^. The presence of obesity is associated with an increase in self-reported bodily pain measures and pain related disability, a decrease in psychological well-being, and reduced physical functioning ^10-12^. The relationship between pain and obesity is likely multi-directional and multi-factorial, including central pain and inflammatory pathways ^13-15^. Provider prescriptions of non-steroidal anti-inflammatory agents used to treat pain are higher amongst obese patients ^16; 17^.

Given the prevalence of obesity in the general and chronic pain populations and its effect on clinical outcomes, it is important to understand how obesity affects the validated pain outcomes commonly used in clinical practice, including pain severity and pain interference. Previous research has elucidated higher pain scores in female, osteoarthritic, and postoperative patients with obesity. However, these studies are often limited by small or homogeneous cohorts, or examine outcomes that are not commonly used by chronic pain providers in a clinical setting ^18-22^. No major study to date has analyzed the impact that obesity has on pain severity and interference scores in a general chronic pain population. Herein, we seek to further explore the association of obesity as a risk factor for pain related outcomes. Outcomes of interest include: pain severity and pain interference. We hypothesized that a higher level of obesity, as measured by body mass index (BMI), would be associated with increased pain severity and pain interference.

## METHODS

### Patient Population

We collected data from a longitudinal observational cohort of chronic pain outpatients seen in the Weill Cornell Medicine (WCM) pain medicine clinic from 7/8/2011 to 10/17/2016. These patients are part of a larger tri-institutional chronic pain database herein referred to as The Registry, which encompasses Pain Services data from WCM, Hospital for Special Surgery and Memorial Sloan-Kettering Cancer Center (MSKCC), New York, New York. The Registry design, enrollment procedures, data collection and analysis have been previously described ^23^. A data sharing agreement exists between WCM and the Health System Innovation and Research Program, University of Utah School of Medicine, Salt Lake City, UT, who maintain the database and assist with the data analysis. Health System Innovation and Research Program merges the coded PRO data in Webcore with data captured in EPIC to form a complete coded limited data set for The Registry ^23^. While patient identifiers are removed, the coded data contains encounter dates, and therefore the data represents a limited data set according to the HIPAA Privacy Rules. Patient data is collected under standard of care and is part of a quality improvement program.

The inclusion criteria for this study included all chronic pain patients, as defined by at least one clinic attendance. If multiple visit had data recorded, the first entry was the value used in analysis. The original exclusion criteria for The Registry was absence of a chronic pain diagnosis, but for this study the exclusion criteria also included patients without BMI data and without a specific type of chronic pain diagnosis for the encounter ^23^.

Patient specific factors and process factors were obtained for each patient at each encounter. These factors included: demographics (Age, Sex, Marital Status, Ethnicity, Language, Religion, and Payor), vitals (height, weight, BP, Temperature, pulse rate), diagnoses, CPT codes, medications, and the survey data which included the pain severity and pain interference scores. BMI was calculated as weight in kilograms divided by the square of height in meters. Participants were categorized into three groups based on BMI: underweight (BMI <18.5), normal weight and overweight (BMI = 18.5-29.9), and obese (BMI ≥30); the normal weight and overweight category was used as the reference in all models.

During each outpatient pain clinic encounter, pain information was collected using the Brief Pain Inventory (BPI) short form. The BPI short form is a validated tool that assesses the presence of pain; pain severity, pain interference, percentage of relief provided by pain treatments or medications (0-100%), and the location of pain at 22 pre-specified sites assessed over a 24-hour recall period ^24^. Patients describe their pain severity on a scale from 0 (“No Pain”) to 10 (“Pain as bad as you can imagine”) when their pain is at its worst, least, on average, and if they are experiencing pain right now^7^. Pain interference is measured on seven daily functions, including general activity, mood, walking ability, normal work (includes both work outside the house and housework), relations with other people, sleep, and enjoyment of life. Each pain interference measure is assessed through a rating scale from 0 (“Does not Interfere”) to 10 (“Completely Interferes”).

### Estimating Opioid Use, Dosage, and Duration

Opioid medications that were being taken and prescribed at each encounter were abstracted from EPIC. For patients included in the Registry, we captured all the opioids recorded for each encounter backward to 11/1/2010 (this date was specified as when clinicians were required to perform medication reconciliation in EPIC and therefore represents the most reliable data).

Resultantly, opioid duration was measured from 11/1/2010 to 9/1/2016, including opioid use prior to entry into the Registry. Based on a discussion with the pain clinicians on patient opioid prescription filling practices, we looked back at least 45 days from their clinic visit for a schedule 2 opioid prescription or refill. We excluded patients receiving intrathecal opioids from this analysis.

### Statistical analysis

Statistical analyses were performed using R Version 3.5.3 (Vienna, Austria)^25^. Bivariate analyses were performed to compare pain outcomes by demographic/clinical patient characteristics as well as to compare demographic/clinical patient characteristics by bmi groups (underweight, normal and overweight, and obese). Bivariate tests included independent two-sample t-tests, Analysis of Variance (ANOVA), or Chi-Squared/Fisher’s Exact tests, as appropriate.Nonparametric equivalents were used for violations of normality.

To examine the effect of BMI on pain severity and pain interference we fit multivariable linear regression models to our data; regression coefficients along with their 95% confidence intervals were reported. We developed separate models for each of the four pain severity scores (worst, least, average, and current) and for total pain interference with the additional inclusion of pain severity as a covariate. All models adjusted for demographic and clinical variables available and considered relevant (age, pain procedure within 45 days prior, opioids prescribed within 45 days prior, and diagnosis). All statistical tests were 2-sided with statistical significance assigned at an alpha level of 0.05.

## RESULTS

A total of 2,509 patients were included in the study. The cohort’s median age was 59 years, median BMI was 27 kg/m^2^, and the most common pain diagnosis category was musculoskeletal pain (77%). About 20% of the patients received a pain procedure within 45 days prior to the pain clinic encounter, and half of the patients (53%) received an opioid prescription within 45 days prior. Differences between BMI groups are shown in Table 1. Some statistical significance represrents underlying differences in the spread of pain scores, as shown through the range. Bivariate tests revealed significant differences between BMI groups for all pain severity scores and for total pain interference score (Table 2).

**Table 1:** Differences Between BMI Groups

|  | BMI <18.5 | BMI 18.5-29.99 | BMI ≥30 | p value | N |
| --- | --- | --- | --- | --- | --- |
| Weight | 106 [98.8;118] | 154 [135;175] | 215 [190;243] | <0.001 | 2509 |
| Height | 65.3 [63.7;69.0] | 66.0 [63.0;69.0] | 65.5 [63.9;69.0] | 0.163 | 2509 |
| Age (continuous) | 52.2 [36.8;69.5] | 59.9 [46.4;71.7] | 59.9 [49.4;68.6] | 0.093 | 2509 |
| Age (categorical) |  |  |  | <0.001 | 2509 |
| 18-44 | 25 (36.8%) | 401 (23.5%) | 120 (16.3%) |  |  |
| 45-64 | 23 (33.8%) | 645 (37.8%) | 364 (49.6%) |  |  |
| 65+ | 20 (29.4%) | 661 (38.7%) | 250 (34.1%) |  |  |
| Pain procedure with 45 days prior |  |  |  | 0.743 | 2509 |
| No | 57 (83.8%) | 1366 (80.0%) | 588 (80.1%) |  |  |
| Yes | 11 (16.2%) | 341 (20.0%) | 146 (19.9%) |  |  |
| Pain procedure with 45 days after |  |  |  | 0.69 | 2509 |
| No | 56 (82.4%) | 1332 (78.0%) | 576 (78.5%) |  |  |
| Yes | 12 (17.6%) | 375 (22.0%) | 158 (21.5%) |  |  |
| Opioid medication within 45 days prior |  |  |  | 0.001 | 2509 |
| No | 25 (36.8%) | 841 (49.3%) | 306 (41.7%) |  |  |
| Yes | 43 (63.2%) | 866 (50.7%) | 428 (58.3%) |  |  |
| IR opioid medications within 45 days prior |  |  |  | <0.001 | 2509 |
| No | 26 (38.2%) | 898 (52.6%) | 321 (43.7%) |  |  |
| Yes | 42 (61.8%) | 809 (47.4%) | 413 (56.3%) |  |  |
| ER opioid medications within 45 days prior |  |  |  | 0.041 | 2509 |
| No | 51 (75.0%) | 1468 (86.0%) | 627 (85.4%) |  |  |
| Yes | 17 (25.0%) | 239 (14.0%) | 107 (14.6%) |  |  |
| Pain Severity and Interference |  |  |  |  |  |
| Worst pain, 24 hours | 8.00 [6.00;9.00] | 8.00 [6.00;9.00] | 8.00 [6.00;9.00] | <0.001 | 2425 |
| Least pain, 24 hours | 5.00 [3.00;7.00] | 4.00 [2.00;6.00] | 5.00 [2.00;7.00] | <0.001 | 2415 |
| Average pain, 24 hours | 6.00 [4.25;8.00] | 6.00 [4.00;7.75] | 6.00 [5.00;8.00] | <0.001 | 2371 |
| Pain now- current | 6.50 [4.00;8.00] | 5.00 [3.00;8.00] | 6.00 [4.00;8.00] | <0.001 | 2370 |
| General Activity | 7.00 [5.00;9.00] | 6.00 [4.00;8.00] | 7.00 [5.00;9.00] | <0.001 | 2345 |
| Mood | 7.00 [4.00;9.00] | 6.00 [3.00;8.00] | 6.00 [3.00;8.00] | 0.001 | 2330 |
| Walking | 6.00 [3.00;9.00] | 5.00 [2.00;8.00] | 7.00 [4.00;9.00] | <0.001 | 2339 |
| Normal work | 7.00 [5.00;9.00] | 6.00 [4.00;9.00] | 7.00 [5.00;9.00] | <0.001 | 2315 |
| Relations with others | 5.00 [1.25;7.00] | 4.00 [0.00;7.00] | 5.00 [0.00;7.00] | 0.014 | 2294 |
| Sleep | 6.50 [3.00;8.00] | 5.00 [2.00;8.00] | 6.00 [3.00;9.00] | 0.002 | 2318 |
| Enjoyment of life | 8.00 [5.00;9.00] | 7.00 [4.00;8.00] | 7.00 [5.00;9.00] | <0.001 | 2297 |
| Interference Total Score | 6.29 [5.00;7.71] | 5.57 [3.29;7.43] | 6.46 [4.14;8.00] | <0.001 | 2359 |
| Diagnosis Category |  |  |  | 0.001 | 2509 |
| Musculoskeletal | 44 (64.7%) | 1298 (76.0%) | 578 (78.7%) |  |  |
| Neuropathic | 8 (11.8%) | 207 (12.1%) | 90 (12.3%) |  |  |
| Other | 11 (16.2%) | 135 (7.91%) | 29 (3.95%) |  |  |
| Unclear/unknown | 5 (7.35%) | 67 (3.93%) | 37 (5.04%) |  |  |

**Table 2:** Multivariable Model Resuslts

|  |  | Pain Severity (Worst 24 Hours) |  | Pain Severity (Least 24 Hours) |  |
| --- | --- | --- | --- | --- | --- |
|  |  | Estimate (95% CI) | P-Value | Estimate (95% CI) | P-Value |
| **BMI** | Underweight | 0.284 (-0.315, 0.884) | 0.353 | 0.667 (-0.049, 1.383) | 0.068 |
|  | ref=Normal |  |  |  |  |
|  | Weight/Overweight |  |  |  |  |
|  | Obese | 0.354 (0.139, 0.569) | 0.001 | 0.556 (0.301, 0.811) | 0.000 |
| **Age** |  | 0.000 (-0.006, 0.006) | 0.936 | -0.002 (-0.009, 0.005) | 0.545 |
| **Pain procedure within 45 days prior** |  | 0.011 (-0.232, 0.253) | 0.93 | -0.157 (-0.445, 0.132) | 0.287 |
| **Opioid prescription within 45 days prior** |  | 0.715 (0.517, 0.914) | <0.001 | 0.655 (0.419, 0.890) | 0.000 |
| **Diagnosis** | ref=Muskuloskeletal |  |  |  |  |
|  | Neuropathic | -0.004 (-0.304, 0.296) | 0.978 | -0.127 (-0.483, 0.230) | 0.486 |
|  | Other | 0.061 (-0.326, 0.448) | 0.758 | -0.491 (-0.949, -0.32) | 0.36 |
|  | Unclear/unknown | -0.290 (-0.764, 0.185) | 0.232 | -0.338 (-0.901, 0.224) | 0.239 |
|  | Worst Pain |  |  |  |  |
|  |  | **Pain Severity (Average 24 Hours)** |  | **Pain Severity (Current)** |  |
|  |  | **Estimate (95% CI)** | **P-Value** | **Estimate (95% CI)** | **P-Value** |
| **BMI** | Underweight | 0.437 (-0.146, 1.020) | 0.142 | 0.748 (0.061, 1.434) | 0.033 |
|  | ref=Normal |  |  |  |  |
|  | Weight/Overweight |  |  |  |  |
|  | Obese | 0.382 (0.171, 0.592) | 0.000 | 0.625 (0.376, 0.874) | 0.25 |
| **Age** |  | 0.003 (-0.003, 0.008) | 0.391 | -0.008 (-0.015, -0.001) | 0.25 |
| **Pain procedure within 45 days prior** |  | -0.118 (-0.358, 0.121) | 0.332 | -0.079 (-0.360, 0.202) | 0.580 |
| **Opioid prescription within 45 days prior** |  | 0.595 (0.400, 0.790) | 0.000 | 0.748 (0.519, 0.978) | 0.000 |
| **Diagnosis** | ref=Muskuloskeletal |  |  |  |  |
|  | Neuropathic | -0.048 (-0.341, 0.245) | 0.748 | -0.104 (-0.450, 0.243) | 0.558 |
|  | Other | -0.149 (-0.530, 0.232) | 0.443 | -0.467 (-0.914, -0.020) | 0.041 |
|  | Unclear/unknown | -0.228 (-0.690, 0.234) | 0.333 | -0.312 (-0.857, 0.234) | 0.263 |
|  | Worst Pain |  |  |  |  |
|  |  | **Pain Interference** |  |  |  |
|  |  | **Estimate (95% CI)** | **P-Value** |  |  |
| **BMI** | Underweight | 0.319 (-0.163, 0.801) | 0.194 |  |  |
|  | ref=Normal |  |  |  |  |
|  | Weight/Overweight |  |  |  |  |
|  | Obese | 0.276 (0.104, 0.449) | 0.002 |  |  |
| **Age** |  | -0.017 (-0.022, -0.013) | 0.000 |  |  |
| **Pain procedure within 45 days prior** |  | -0.054 (-0.248, 0.141) | 0.588 |  |  |
| **Opioid prescription within 45 days prior** |  | 0.425 (0.263, 0.586) | 0.000 |  |  |
| **Diagnosis** | ref=Muskuloskeletal |  |  |  |  |
|  | Neuropathic | -0.151 (-0.392, 0.091) | 0.222 |  |  |
|  | Other | -0.342 (-0.080, 0.681) | 0.030 |  |  |
|  | Unclear/unknown | 0.300 (0.657, 0.721) | 0.122 |  |  |
|  | Worst Pain | 0.689 (0.657, 0.721) | 0.000 |  |  |

For each multivariable model looking at pain severity (as either the worst, least, average, or current pain within the past 24 hours), obese patients demonstrated significantly higher scores than normal weight patients after controlling for age, pain procedure within the 45 days prior to the pain clinic encounter, opioid prescription within the 45 days prior, and diagnosis category. Specifically, obese patients had an average 0.354 higher severity score at their worst pain within the past 24 hours, 0.556 at their least pain within the past 24 hours, 0.382 at their average pain in the past 24 hours, and 0.625 at their current pain than normal weight patients. Similarly, for the multivariable model looking at overall pain interference, obese patients demonstrated significantly higher scores than normal weight patients after controlling for the same factors in addition to pain severity (as measured as the worst pain in the past 24 hours). Specifically, obese patients had an average 0.276 higher overall pain interference score than normal weight patients.

Confirming our assumption, pain severity (as measured as the worst pain in the past 24 hours) was significantly associated with overall pain interference; specifically, overall pain interference increased on average by 0.689 points for each additional point higher in worst pain severity. An additional result of note is that underweight patients also showed a significant difference compared to normal weight in the pain now severity model (p=0.033).

Opioid use was statistically significant for each pain severity and pain interference outcome used in multivariable modeling (p<0.0001 for all outcomes). 58.3% of obese patients and 63.2% of underweight patients were prescribed opioid medications within 45 days prior compared to only 50.7% of normal weight patients (p=0.001), with all BMI groups being more likely to take pain medications than not. Differences between BMI groups also existed when looking specifically at IR opioid medications and ER opioid medications prescribed. 56.3% of obese patients and 61.8% of underweight patients were prescribed IR opioid medications compared to 47.4% of normal/overweight patients (p <0.001). About 14% of both obese and normal/overweight patients were prescribed ER opioid medications compared to 25% of underweight patients (p=0.041).

## DISCUSSION

### Pain Prevelance

Our study showed that pain clinic patients with obesity experience higher pain severity and pain interference scores than normal weight and overweight patients. Our findings corroborate prior literature that shows that obesity is a risk factor for chronic pain ^26-29^, and demonstrates that obesity is an independent risk factor for experiencing more pain severity and pain that may interfere with patients’ daily function. Heim et al. reported that in a population of older adults, obese patients were more likely to develop pain, and that the frequency of pain increased more drastically with increased BMI, especially for women ^30^. Their study found that, regardless of weight changes, someone with a higher BMI is more likely to longitudinally develop chronic pain ^30^.

### Pain Severity

Our study found that obesity was related to pain severity. Patients’ rating of worst pain in the last 24 hours was statistically significant, with increased pain severity found in obese patients compared to the normal and over-weight patients. Obesity as a risk factor for pain severity has been found in many other studies, ranging from adolescents to geriatric populations ^31; 32^. Chronic pain development in an obese population, particularly morbidly obese (BMI>35), after a motor vehicle accident had a significant effect on increasing pain severity during multiple follow-up visits within a year ^22^. However, some studies have disputed the relationship between pain severity and obesity ^19; 33^. Taspinar et al. did not find obesity to influence pain severity for university students with nonspecific back pain. However, this study included only university students, which limits its generalizability to the wider chronic pain population, which includes patients of a larger age range.

Looking at the underweight population, the literature agrees with our finding of increased current pain severity in the underweight population compared to the normal group ^34^. Previously, this has also influenced the prescription of opioid medications for pain relief, specifically in cancer populations, but the true clinical relevance has yet to be determined.

### Pain Interference

Pain severity is related to pain interference, as it is not unexpected that higher levels of pain intensity would have more drastic effects on functioning ^5^. In our study, pain interference scores increased with increased ratings of worst pain in the last 24 hours. Pain severity and frequency has been shown to directly predict pain interference and quality of life ^35-37^. Of note, our findings contradict with several studies that did not find an association between pain severity and pain interference ^19^. This finding could be due to a threshold effect that occurs where low levels of pain intensity are not associated with functional interference and where higher levels of pain intensity increase the probability of higher levels of pain interference in a nonlinear fashion ^5-7^.

In our study, obese patients had higher pain interference than normal weight patients.This follows previous trends finding that obese patients have a higher frequency of pain interference and more severe disability from pain ^19; 29; 32; 36^. Only one study found obesity to be independent of pain interference scores ^18^.

While these findings are statistically significant, clinical implication has yet to be determined within this research. Previous literature has speculated that at least a 30% change from baseline on the BPI is required for an impacted quality of life ^38-40^.

### Opioid Use

Pain experience is also altered by opioid use, which is commonly prescribed for pain management. Opioid usage was statistically significant between the three BMI categories for both IR and ER medication prescription 45 days prior to the visit. IR medication prescription was more commonly used within all three BMI groups compared to ER medication, with underweight and overweight patients having increased prescriptions compared to the normal weight group. In multivariable modelling, opioid usage was significantly associated with higher pain severity and pain interference.The increase in pain severity and interference increasing with opioid use is unique to our study. A previous study found that opioid medications had no statistically significant difference between BMI (obese or not obese), but opioid use did relate to an increase in pain interference ^18^. Provider prescriptions of non-steroidal anti-inflammatory drugs used to treat pain found that they are higher amongst obese patients ^16; 17^. Similar to our studies, previous research analyzing the use of opioids and obesity within the context of pain are often limited in a cross-sectional setting due to the unknown timeline of pain characteristics and opioid prescriptions, thus only being able to examine correlation and not causation.

### Limitations

Our study is limited by retrospective data, multi-institutional data collected primarily from tertiary care centers, and the high prevalence of musculoskeletal pain in our population. Seventy-seven percent of all participants in this study had a diagnosis of musculoskeletal pain, and 79% of obese participants diagnosed with that as well, being only slightly higher than the normal and overweight category (76.0%) and significantly higher than the underweight group (64.7%). Previous literature often restricted cohort populations to those with specific types of pain, like musculoskeletal, since its development has been more easily associated with obesity ^31; 32; 41^. Musculoskeletal pain has also been connected with more pain locations, which is further increased within an obese population ^32; 41^. While a diagnosis of musculoskeletal pain may influence obesity and pain severity scores, it is an extremely prevalent pain diagnosis with clinical implications in both our population and the general population, and thus could not be ignored.

Our study was also limited by the categorization of weight. For our study, obesity was defined as anyone with a BMI greater than or equal to 30. Further stratification of this obese category has shown trends that indicate greater pain in the severely obese. For McCarthy et al., the severely obese, or those with a BMI of at least 35, were associated with four times the probability of having chronic pain compared to normal patients, whereas those with BMI=30-34.99 were only twice as likely ^32^. This stratification more drastically shows the impact that obesity poses on the experience of chronic pain. Deyo et al. also showed that for those in the top 20%, the prevalence of pain has been found to be significantly higher than those in the bottom 20% of the obese category ^28^.

Our study only included patients that visited the outpatient pain clinics included in The Registry at least once with BMI information and a diagnosis for the encounter. The survey data analyzed was that of the first survey in the registry for each patient. While the first available BMI value was used as well, this did not consistently align with the first full survey dataset since patients often had multiple pain clinic encounters. By identifying patients retrospectively, we weren’t able to follow their progress over more visits longitudinally. In addition, while The Registry is a multi-site database, it gathers information solely from New York City-based sites, which could limit generalizability of our results.

In addition, our study was not able to capture the prevalence of psychiatric comorbidities within our chronic pain population. Many other studies have found that this dramatically impacts both obesity and pain, often increasing the likelihood or severity of both ^9-11; 26; 27; 32; 42; 43^. Further research is warranted to assess the role of psychiatric comorbidities on pain interference and severity within a chronic pain population, especially as it may influence management choices and clinical prognosis.

## Conclusions

Our study demonstrated that pain severity and interference is impacted by obesity and opioid use. The management of chronic pain is multifaceted and complex, particularly in the obese chronic pain population. Our study suggests that these patients’ chronic pain is characterized not only by higher pain severity, but also greater interference with activities of daily living. Our findings suggest that counseling patients about their BMI, health maintenance, and setting reasonable expectations in regards to physical functioning and prognosis may improve their pain management treatment in terms of lessening their pain severity and pain interference. Further prospective research is warranted to understand the relationship between reducing BMI and pain interference scores.

## Data Availability

Data are available upon request.

## ACKNOWLEDGEMENTS

We would like to thank the Weill Cornell Medicine Department of Anesthesiology for their support.

## Notes

**Conflicts of Interest:** Charles Inturrisi receives compensation as described in a 2017 license agreement between Charles E.. Inturrisi (and Paolo Manfredi) and Relmada Therapeutics, Inc.. for the development of compounds (including d-methadone) for the treatment of depression. Otherwise, the authors declare no conflicts of interest.

### Competing Interest Statement

Charles Inturrisi receives compensation as described in a 2017 license agreement between Charles E.. Inturrisi (and Paolo Manfredi) and Relmada Therapeutics, Inc.. for the development of compounds (including d-methadone) for the treatment of depression. Otherwise, the authors declare no conflicts of interest.

### Funding Statement

The authors have no sources of funding to declare for this manuscript.

